# What influences ambulance clinician decisions to pre-alert Emergency Departments: a qualitative exploration of decision-making in three UK Ambulance Services

**DOI:** 10.1101/2023.12.14.23299973

**Authors:** Rachel O’Hara, Fiona Sampson, Jaqui Long, Joanne Coster, Richard Pilbery

## Abstract

**Background:** Ambulance clinicians use pre-alerts to inform receiving hospitals of the imminent arrival of a time-critical patient considered to require immediate attention, enabling the receiving Emergency Department or other clinical area to prepare. Pre-alerts are key to ensuring immediate access to appropriate care, but unnecessary pre-alerts can divert resources from other patients and fuel ‘pre-alert fatigue’ amongst ED staff. This research aims to provide a better understanding of pre-alert decision-making practice.

**Methods:** Semi-structured interviews were conducted with 34 ambulance clinicians from three ambulance services and 40 ED staff from six receiving EDs. Observation (162 hours) of responses to pre-alerts (n=143, call-to-handover) was also conducted in the six EDs. Interview transcripts and observation notes were imported into NVIVO and analysed using thematic analysis.

**Findings:** Pre-alert decisions involve rapid assessment of clinical risk based on physiological observations, clinical judgement, and perceived risk of deterioration, with reference to pre-alert guidance. Clinical experience (pattern recognition and intuition) and confidence helped ambulance clinicians to understand which patients required immediate ED care upon arrival or were at highest risk of deterioration. Ambulance clinicians primarily learned to pre-alert ‘on the job’ and via informal feedback mechanisms, including the ED response to previous pre-alerts. Availability and access to clinical decision support was variable, and clinicians balanced the use of guidance and protocols with concerns about retention of clinical judgement and autonomy. Differences in pre-alert criteria between ambulance services and EDs created difficulties in deciding whether to pre-alert and was particularly challenging for less experienced clinicians.

**Conclusion:** We identified potentially avoidable variation in decision-making, which has implications for patient care and emergency care resources, and can create tension between the services. Consistency in practice may be improved by greater standardisation of guidance and protocols, training and access to performance feedback, and cross-service collaboration to minimise potential sources of tension.

## BACKGROUND

Ambulance clinicians use pre-alert calls to inform receiving Emergency Departments (EDs) of the imminent arrival of a time-critical patient considered to require immediate attention. UK national guidance states that pre-alert calls ‘should be used to provide information about the patient that will enable the receiving ED or other clinical area to prepare a different or special response’[p1] ^1^ Preparation can include assembling staff from other areas of the hospital (e.g. trauma team), preparing specialist equipment and allocating space within the resuscitation area. ^2^

Pre-alerts can facilitate faster access to ED care, improved processes and better clinical outcomes for time-critical patients, ^3 4^ but unnecessary pre-alerts can divert resources from other patients and fuel ‘pre-alert fatigue’ amongst ED staff. ^5^ Pre-alert fatigue may contribute to ED staff losing trust in pre-alert calls, a potential risk for patients and source of frustration for ambulance clinicians that they are not taken seriously.

Significant variation was found in guidance between UK ambulance services on the clinical criteria that should trigger a pre-alert,^6^ which is likely to be a factor influencing differences in pre-alert practice. For example, studies have reported that an estimated 13%-28% of patients eligible for pre-alert had not received one and 42%-56% of pre-alerted patients did not meet the established criteria. ^3 4 7^ Even where there are clear guidelines for pre-alert (e.g. stroke, trauma)^8^, pre-alerts are only used in a proportion of cases. ^9^ Similarly, Sheppard et al. identified disparities in how ambulance clinicians and receiving ED staff interpreted protocols and the need to pre-alert for suspected stroke, leading to frustration for ambulance and ED staff.^7^

Variation in pre-alert practice is a concern regarding patient safety^10^ and potential inequities in care, ^11^ therefore, it is important to understand the possible contributory factors. Although joint UK Ambulance Service and ED guidance was developed in 2020^1^, there is a lack of evidence on pre-alert practice. This paper aims to provide a better understanding of ambulance clinicians’ pre-alert decision-making practice.

## METHODS

### Design

This research was problem-driven and a qualitative approach was employed to explore pre-alert decision-making by ambulance clinicians. The methods included interviews with ambulance crew/clinicians and ED staff, and non-participant-observation within receiving EDs.

Ethical approval for the research was obtained from NHS ethics North East - Newcastle & North Tyneside 2 Research Ethics Committee (Ref: 21/NE/0132).

### Settings

Three ambulance services in England were selected on the basis of their interest in the research and contributed to the design. EDs included one Major Trauma Centre (MTC) and one Trauma Unit (TU) in each ambulance service region that were identified as having sufficiently high numbers of pre-alerts for observations.

### Data collection

Semi-structured telephone or online interviews were conducted by JL and JC with a purposive sample^12^ of 34 ambulance clinicians from three ambulance services and 40 ED staff from six linked receiving EDs. Ambulance Service interviewees were recruited via open communications targeting ambulance crews. ED staff were recruited via direct invitation during observations and via local research leads who invited staff in particular roles to take part. This helped to ensure that a range of roles at each site were represented (e.g. senior, junior, medical, nursing). Interviewees were provided with study information and completed a consent form. Interviews were audio-recorded and transcribed verbatim. The interview topics are presented in box 1.

#### Box 1

Interview topics - ambulance service and ED

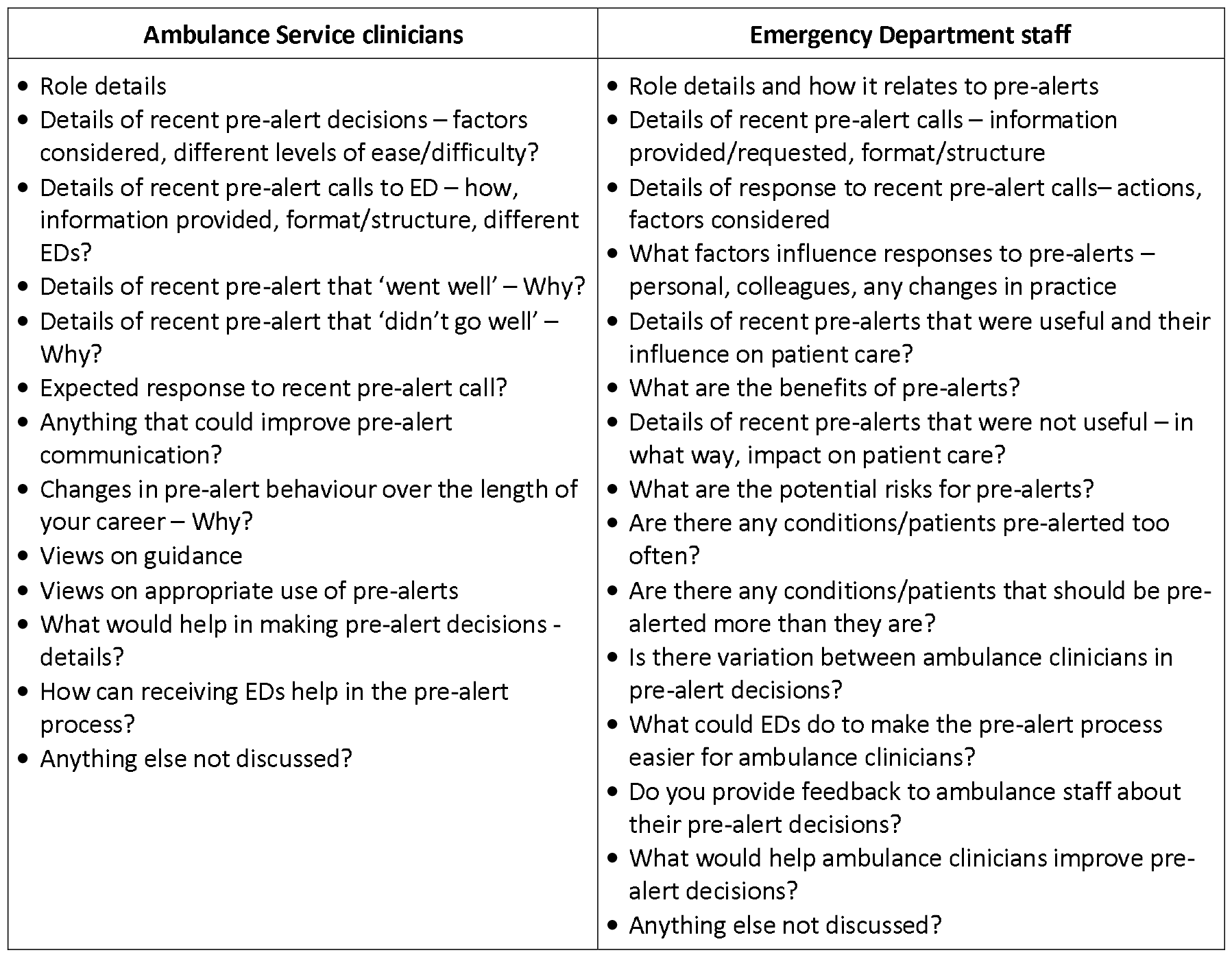

Box 1: Interview topics - ambulance service and ED
| Ambulance Service clinicians | Emergency Department staff |
| --- | --- |
| • Role details<br>• Details of recent pre-alert decisions – factors considered, different levels of ease/difficulty?<br>• Details of recent pre-alert calls to ED – how, information provided, format/structure, different EDs?<br>• Details of recent pre-alert that ‘went well’ – Why?<br>• Details of recent pre-alert that ‘didn’t go well’ – Why?<br>• Expected response to recent pre-alert call?<br>• Anything that could improve pre-alert communication?<br>• Changes in pre-alert behaviour over the length of your career – Why?<br>• Views on guidance<br>• Views on appropriate use of pre-alerts<br>• What would help in making pre-alert decisions - details?<br>• How can receiving EDs help in the pre-alert process?<br>• Anything else not discussed? | • Role details and how it relates to pre-alerts<br>• Details of recent pre-alert calls – information provided/requested, format/structure<br>• Details of response to recent pre-alert calls– actions, factors considered<br>• What factors influence responses to pre-alerts – personal, colleagues, any changes in practice<br>• Details of recent pre-alerts that were useful and their influence on patient care?<br>• What are the benefits of pre-alerts?<br>• Details of recent pre-alerts that were not useful – in what way, impact on patient care?<br>• What are the potential risks for pre-alerts?<br>• Are there any conditions/patients pre-alerted too often?<br>• Are there any conditions/patients that should be pre-alerted more than they are?<br>• Is there variation between ambulance clinicians in pre-alert decisions?<br>• What could EDs do to make the pre-alert process easier for ambulance clinicians?<br>• Do you provide feedback to ambulance staff about their pre-alert decisions?<br>• What would help ambulance clinicians improve pre-alert decisions?<br>• Anything else not discussed? |

Observation of ED staff responses to pre-alert calls (n=143) and patient handover in resus was conducted in the six EDs, and included informal conversations with hospital and ambulance staff present. Observations were undertaken between June 2022-April 2023 by JL and JC. Initial familiarisation observations involved the research lead (FS) to clarify the scope of data collection. A total of 25 observation sessions were undertaken (162) hours. For some sessions (n=9; 39 hours) both researchers attended together to ensure continuous cover and compare observations. Initial site visits communicated to staff the purpose of the research and what it involved. Additional communication to inform staff of the observations included emails, posters, leaflets, attendance at briefings and researcher introductions during the actual observations. Staff were offered the chance to opt-out if they did not wish to be observed (nobody opted-out). Handwritten and audio-recorded field notes including observations, informal conversations and researcher reflections, were fully transcribed.

### Details of interview participants and observations

Details of ambulance clinician interview participants are provided in Table 1 and ED staff participants are provided in Table 2. Details of ED observations are provided in table 3.

**Table 1:**
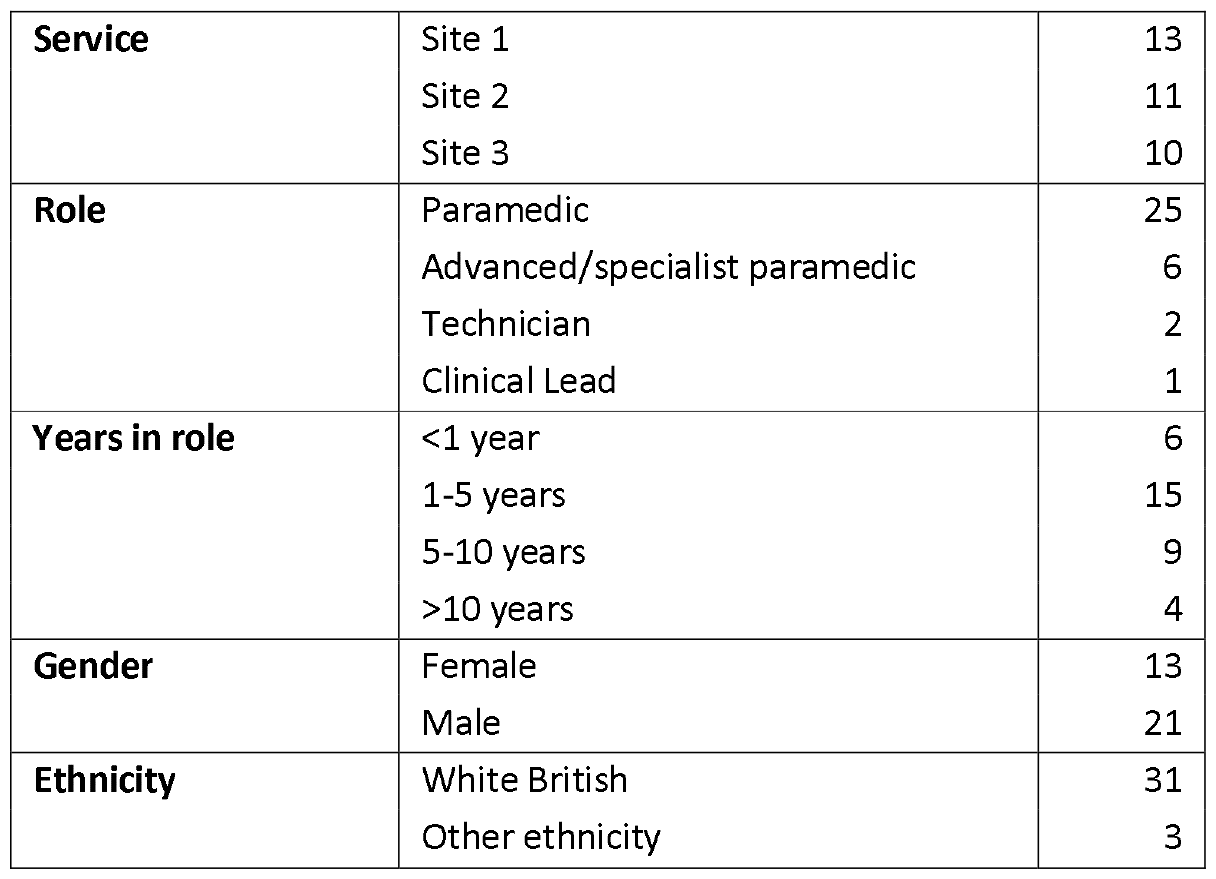
Details and number of participants per ambulance service.

**Table 2:**
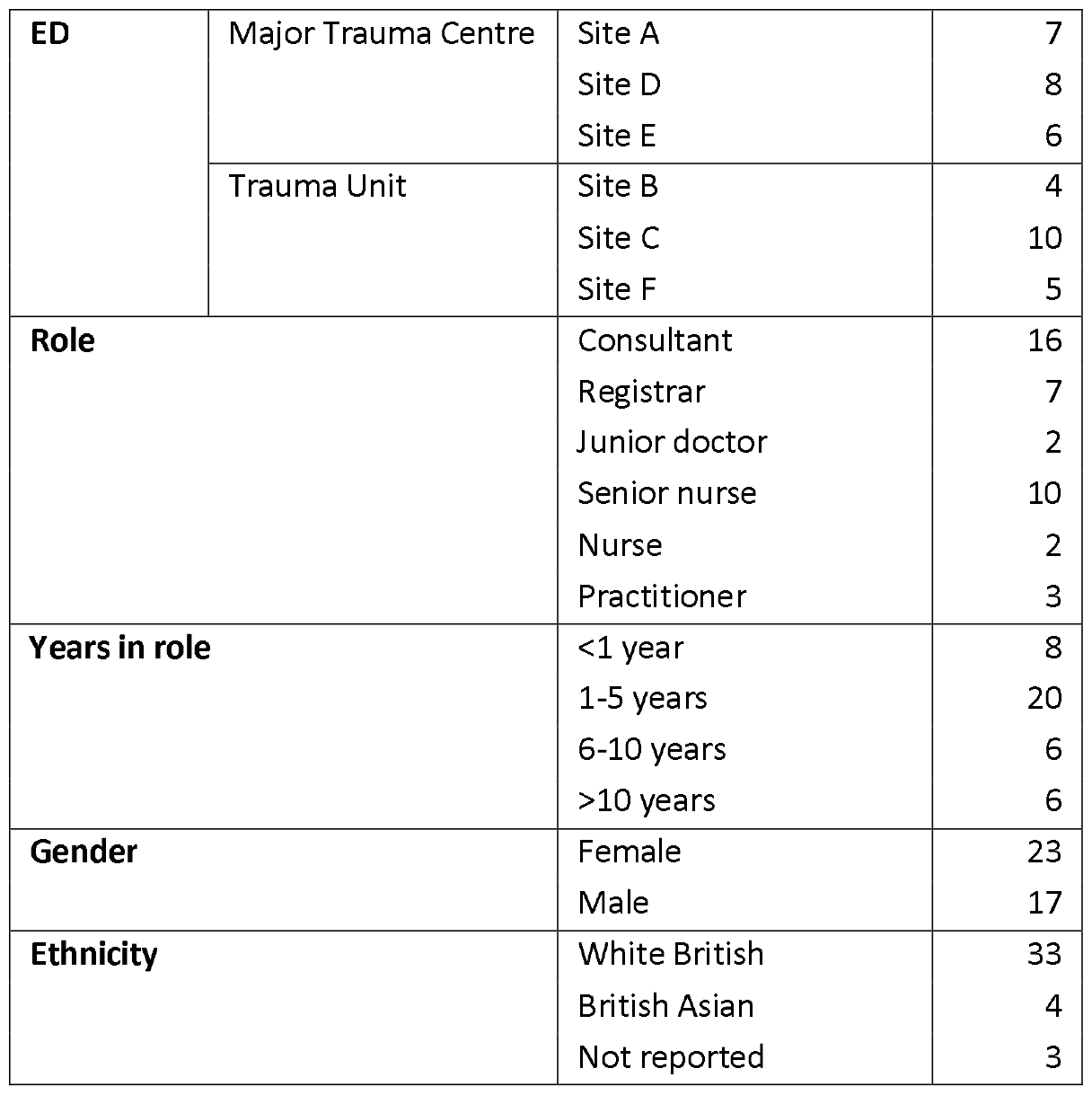
Details and number of staff participants per ED.

|  |  |  |  |
| --- | --- | --- | --- |
| ED | Major Trauma Centre | Site A | 7 |
|  |  | Site D | 8 |
|  |  | Site E | 6 |
|  | Trauma Unit | Site B | 4 |
|  |  | Site C | 10 |
|  |  | Site F | 5 |
| Role |  | Consultant | 16 |
|  |  | Registrar | 7 |
|  |  | Junior doctor | 2 |
|  |  | Senior nurse | 10 |
|  |  | Nurse | 2 |
|  |  | Practitioner | 3 |
| Years in role |  | <1 year | 8 |
|  |  | 1-5 years | 20 |
|  |  | 6-10 years | 6 |
|  |  | >10 years | 6 |
| Gender |  | Female | 23 |
|  |  | Male | 17 |
| Ethnicity |  | White British | 33 |
|  |  | British Asian | 4 |
|  |  | Not reported | 3 |

**Table 3:**
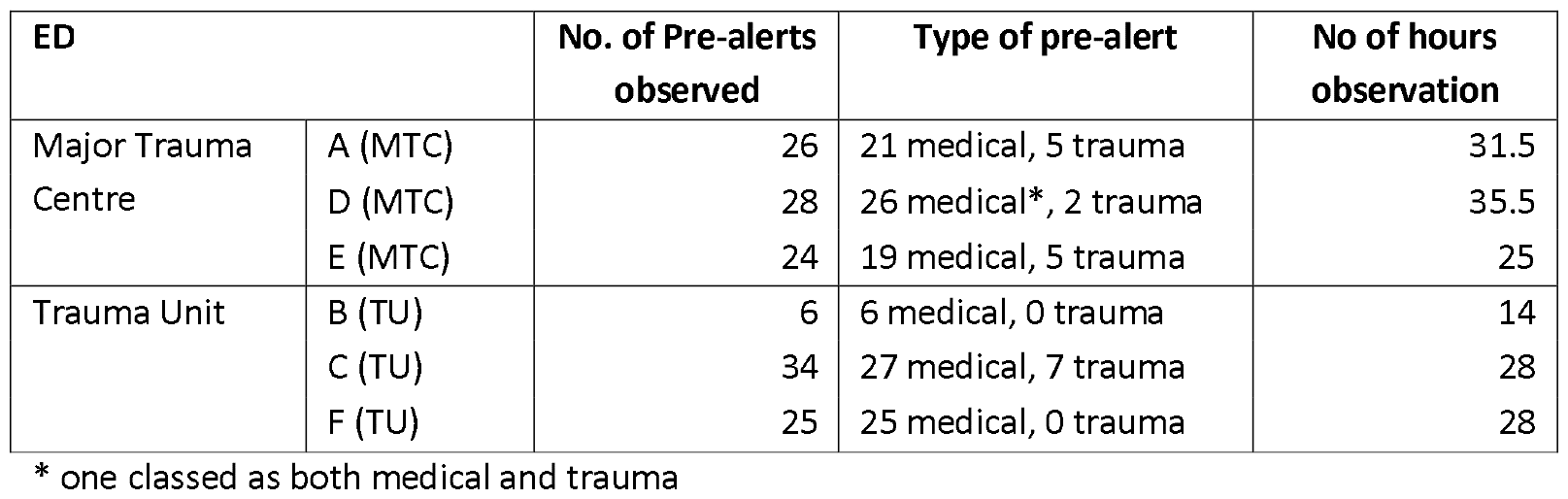
Details of pre-alerts observed.

### Data analysis

Interview transcripts and observation notes were imported into NVIVO^13^ and analysed using thematic analysis. Thematic analysis is an iterative process for systematically analysing qualitative data, it is useful in identifying similarities and differences across data sources. ^14^ An initial coding framework was developed by RO, JL, JC and FS. Coding involved systematically reviewing and coding all data by RO (who had not undertaken any fieldwork) and JL (who undertook the majority of data collection), after agreement on interpretation of the codes. Subsequent code changes were discussed and documented. Further analysis by the four researchers involved reviewing codes to identify and refine themes, and regular discussion of the emerging findings.

The researchers were all non-clinical health service researchers with social science/psychology backgrounds and 8-30 years qualitative research experience.

### Patient and Public involvement

The study Patient and Public Involvement (PPI) group included people with lived experience of pre-alerts, either through being pre-alerted themselves or as a carer or family member of someone who has been pre-alerted. A member of the PPI group was a co-applicant and attended project management meetings. Additional regular PPI group meetings were held to discuss progress and a workshop was conducted to share the findings and elicit feedback.

## FINDINGS

We identified five key aspects of ambulance crew decision-making that affected pre-alert practice: assessing clinical risk to identify patients for pre-alert, clinical experience and confidence, learning and developing good practice, access to clinical decision support, and differences between receiving EDs. Illustrative interview quotes are presented in italics.

### Making judgments about the need for pre-alert based on immediate and potential clinical risk

Pre-alert decisions involve a rapid assessment of clinical risk to identify those patients in need of immediate attention at hospital (e.g. access to a space in resus, with specialist staff and equipment ready). Assessments were based upon physiological observations, clinical judgement, and perceived risk of deterioration, taking account of guidance specifying which patients need to be pre-alerted. Additional considerations that could modify a risk assessment based on their immediate presentation included, pre-existing conditions or past medical history, either due to higher ‘normal’ observations (e.g. respiratory rate for patients with COPD) or the potential for deterioration.

*‘When you’ve got somebody who’s – for example, a heart attack, it’s a no brainer. Then you’ve got things such as working with sepsis tools where it’s very, very “Shall we? Shan’t we?” – there’s other times, with 15 years’ experience, where the numbers might look right but the patient doesn’t’ [AS17_Paramedic]*

*‘It’s not just how they present and how their observations are, it’s also about which direction the observations are going in*.*’ [AS21_Paramedic]*

*‘Any breathing difficulties you think - well that needs a pre-alert*…*But there are plenty of patients that have pre-existing conditions, and it’s an exacerbation of that. Such as COPD, Chronic Obstructive Pulmonary Disease, where even though they are quite poorly-seeming, actually it’s not that different from their baseline*.*’ [AS1_Emergency Medical Technician]*

Certain clinical conditions were acknowledged as clearly necessitating a pre-alert for time critical intervention (e.g. ST-elevation myocardial infarction-STEMI, stroke, major trauma). Medical pre-alerts were often less obvious and based on clinical observations (e.g. heart/respiratory rate, blood pressure, temperature), but the patient’s appearance alone, could give cause for concern. Clinicians commented that decisions were more difficult where observations were within relatively normal range, but the patient looked severely ill as they will have to justify a pre-alert to the receiving ED.

*‘You do the ECG and it’s normal – something is telling you that something is not quite right. I think they’re the more tricky ones, because you’re trying to convince the hospital that you need something different … so I think it’s just about having that confidence that if something is not right then just make the call*.*’ [AS41_Specialist Paramedic]*

*‘I think a lot of the time the ambulances are quite good at just phoning up and saying yeah they don’t trigger [pre-alert criteria] but they just look awful*.*’ [ED40_Registrar]*

Children triggered a higher degree of concern and increased likelihood of pre-alert, partly due to concerns about the risk of sudden deterioration, but also clinicians limited experience with this patient group. ED staff noted that elderly trauma patients generate relatively less concern as ambulance clinicians possibly underestimate the risk associated with falls.

*The grey areas in every way shape or form - children. We worry, and rightly so, we worry a lot about children, before we’ve examined them properly*.*’ [AS17_Paramedic]*

*‘I think a lot of paediatrics is over alerted, but I think that’s purely because of the emotiveness of the scene and the experience of the clinicians seeing the children’ [ED30_Consultant]*

*‘The older adults with trauma…there’s been an under appreciation that they’ve got significant injuries… there is a massive under use of that [silver trauma] tool… So then I think there is a delay for them to be seen by senior decision makers, doctor, a delay to imaging, delay to getting off scoop, or having their head immobilised. And laying flat for an older patient is not good*.*’ [ED43_Consultant]*

### The role of clinical experience and confidence in pre-alert decision-making

Experience and confidence were identified as key factors in decision-making. Ambulance clinicians used clinical experience (pattern recognition and intuition) to make judgements based on previous exposure to specific clinical presentations and conditions. This enabled them to differentiate between critical and less obviously critical patients, and potential deterioration.

*‘By experience you’ve seen the same thing a lot of times and you can perhaps see a pattern where you know what’s going to happen, it just hasn’t happened yet*.*’ [AS17_Paramedic]*

*‘Subconsciously, you recognise things differently, the more experience you get and the more knowledge that you gain*.*’ [AS4_Paramedic]*

*‘As I’ve gone through my career as a technician, you pick up more while you’re on the road… I maybe pre-alert a bit more than what I maybe did right at the beginning. For example when I did my tech training seven years ago, aortic dissection was unheard of*.*’ [AS43_Emergency Medical Technician]*

Clinicians regarded pre-alerts as necessary when they were unable to stabilise the patient, which varied according to their role-related knowledge and skills. Higher level clinicians were able to deliver treatments to stabilise patients or reduce the risk of deterioration.

*‘Am I happy that my patient is stable enough that I can look after them or do I want sort of sooner intervention?’ [AS16_Paramedic]*

*‘Supra-ventricular-tachycardia…they can need medications that we can’t give, that they need at hospital…equally it’s a long term condition they suffer with and they aren’t otherwise compromised*.

*So that can be a bit of a judgement call, not everyone would pre-alert’ [AS1_Emergency Medical Technician]*

*‘The more experienced paramedics that’d be less likely to pre-alert because they’d feel more comfortable managing the patients themselves and aren’t so worried about things’ [ED25_Consultant]*

UK ambulance crews comprise different combinations of roles (e.g. technician, paramedic, advanced paramedic) according to local staffing models, which was identified by ED clinicians as a factor in pre-alert variation between different ambulance services. Lower skilled clinicians and private crews were perceived as pre-alerting more.

*‘I’d say 98% of the time, that is our gold standard, we will have a paramedic on every vehicle’ [AS15_Paramedic]*

*‘We’re very much technician led…I’m usually with a fellow technician or sometimes trainee technician or emergency care assistant’. [AS43_Emergency Medical Technician]*

*‘There’s always going to be a variation in individual practice but there is notable variation between ambulance services. Pre-alerts from non-regular paramedic teams or tech crews are less likely to match what they then bring in’. [ED56_Consultant]*

Experience was identified as affecting confidence and risk tolerance. Whilst newly qualified clinicians were generally regarded as more risk-averse, their lack of exposure to infrequent high acuity conditions could also lead to over-confidence. Concerns were raised that delays at hospitals limit the exposure of newly qualified and trainee clinicians to different patients, affecting their pre-alert experience and confidence. The risk of under-alerting by less experienced clinicians was raised as an issue by ED staff due to large numbers of newly qualified ambulance clinicians.

*‘I’m still quite new to it, so…probably, as a clinician, err on the side of caution a lot more than more experienced people’ [AS11_Newly Qualified Paramedic]*

*‘When I first qualified I was probably more cautious. Whereas now obviously six years in you just get a feeling for things more. And I think that I’m more confident now in saying this patient’s stable’ [AS31_Paramedic]*

*‘I think certainly the experience levels of paramedics generally has changed and I think there are a lot more junior paramedics’ [ED36_Consultant]*

Ambulance crew mentioned the importance of being able to justify their decisions and documenting the reasons to safeguard themselves personally and professionally. They attributed risk-averse decisions to perceptions of a ‘blame culture’ within their organisation and a lack of confidence that they would be supported if something went wrong. This could be mitigated by calling the ED for advice, which provided reassurance that concerns were shared. ED staff also commented on apparent defensive decision-making by ambulance clinicians but understood the reasons for this.

*‘I didn’t think it needed pre-alerting either, but ultimately if I don’t, I’m going against procedure, and if anything goes wrong, where do I stand there? leaves not a lot of leeway for clinical judgement*

*‘[AS28_Paramedic]*

*‘If we get pre-alerted about a kid, from our perception of it is we think what have they rung that through for…if they hadn’t have rung it through and they missed it, and there was something wrong, then they’d be the first people to be criticised and probably hung out to dry about it. As much as we probably whinge about being pre-alerted, probably better to be pre-alerted*.*’ [ED23_Consultant]*

*‘The paramedics are often very worried about making the wrong decision, and so will telephone to see what we think…that’s not always that helpful, and is not what the process is supposed to be. But, they’re in a tough situation and they are often terrified of making the wrong decision*.*’ [ED5_Consultant]*

### Learning and developing good decision-making practice

Ambulance clinicians generally learned to pre-alert ‘on the job’ via a mentor or other experienced clinicians rather than formal training. Some ambulance clinicians had accessed resources for personal learning (e.g. podcasts) to understand how to undertake pre-alerts but most expressed a desire for better training. Clinicians from both ED and ambulance services highlighted the need for opportunities to enhance their understanding of practice in the other setting, for example, placements and joint events to share perspectives on pre-alert practice.

*‘A lot of it comes from my experience as a student, seeing what other paramedics did, and speaking to other paramedics…So that is largely where my decisions come from*.*’ [AS5_ Newly Qualified Paramedic]*

*‘I had a really good paramedic mentor who had a lot of clinical experience…I think I’ve stayed in that method*.*’ [AS12_ Newly Qualified Paramedic]*

*‘Just more training and engagement and maybe just even hospital staff coming into ambulance stations or us having joint training with the staff and saying what do you do, what do you need from me’ [AS24_Paramedic]*

Feedback on pre-alert outcomes was regarded as key to informing future practice and developing confidence by reassuring clinicians about the appropriateness of decisions, rather than making assumptions based on the ED response experienced or observed. However, both ambulance and ED clinicians discussed the challenges of sharing feedback, particularly in the context of extremely busy and pressured environments.

*‘I think sometimes hospitals can make it difficult, because you want to alert something in, and then they turn round and go what are you alerting this in for. But then, you don’t alert the same thing in next time and somebody else tells you something different…I don’t really care to be honest, I think if they’re sick, they are being alerted in*.*’ [AS18_Paramedic]*

*‘When clinicians that have been in the service longer sometimes say you don’t need to pre-alert. The argument I put forward is it’s easier to step someone down from resus rather than have to escalate upwards…this is what I’ve always been told by ED staff’ [AS7_Paramedic]*

*‘You often hear crews “ooh last time I got told off for not calling this through”, or “last time I did call this through and I got in trouble”. So you do hear a lot of that, which I think’s a bit of a shame really. But perhaps that negative behaviour, in their reception before, has sort of influenced their ongoing practice*.*’ [ED2_Consultant]*

*‘We have a tendency to sort of roll our eyes at some of the paramedic decisions. But, I think it is really, really tough for those guys cos the high acuity stuff they get to see very infrequently’ [ED5_Consultant]*

Limited use of formal systems for feedback was identified (e.g. patient QR codes and request cards), but most feedback occurred informally via clinician interactions at handover or later that day. On some occasions, a paramedic located in the ED acting as a Hospital Ambulance Liaison Officer (HALO) to support the management of ambulance patients, was able to facilitate communication and feedback.

*‘Sometimes I might have a word with the HALO officer and just go “By the way, can you make sure they know X, Y and Z for next time”*.*’ [ED14_Consultant]*

Ambulance clinicians expressed a desire for individual-level and organisational-level feedback on pre-alert practice to support learning and development.

*‘I think we need some regional shared learning and training. I think that would be the way. Look at areas of really good practice and really poor practice’ [AS29_Specialist Paramedic]*

### Access to clinical decision support

Ambulance clinicians identified a range of sources of decision support, including guidance, protocols, decision tools (e.g. major trauma tool, sepsis tool), and telephone support. For some conditions, pre-alert recommendations are built into treatment protocols available via an app, which provides clinical guidelines for UK paramedics.^15^ Protocols and decision tools were considered valuable in decision-making, particularly for less experienced clinicians, potentially reducing decision variation and the risk of under-alerting. However, ambulance clinicians highlighted concerns, including increased time taken to make a decision and the inability to take contextual factors into account (e.g. underlying conditions).

*‘I didn’t actually feel that this person needed resus, the only reason I was pre-alerting was because we have guidance that says they needed pre-alerting*.*’ [AS5_ Newly Qualified Paramedic]*

*‘If you’re taking 20 minutes to make a clinical decision because you’re going through all this criteria, that’s 20 minutes of dilly dallying about maybe on scene’ [AS4_Paramedic]*

There was concern about the retention of clinical judgement and autonomy, particularly where patients might not meet certain thresholds but appear very ill and are judged to need pre-alerting. Where patients met pre-defined pre-alert criteria but were not considered to be time-critical, clinicians indicated they would generally pre-alert but inform the ED of their assessment. Some viewed this as reducing their personal responsibility, whereas others felt more confident to exercise their clinical judgement over the need to pre-alert.

*‘Going on the major trauma tool I didn’t need to alert the receiving hospital, however using my judgement of this…he would be (a) inappropriate to hold outside A&E especially with his injuries and (b) he’s going to need a trauma CT, trauma survey…I’ve always thought anyway there needs to be an element of being an autonomous paramedic rather than just following a flow chart in front of you*.*’ [AS7_Paramedic]*

*‘Now it doesn’t really allow for a competent experienced clinician to use their judgement’ [AS20_Paramedic]*

Ambulance clinicians generally valued telephone advice and support, preferably timely and provided by clinicians with more expertise than them (e.g. doctor, nurse specialist, advanced paramedic), acknowledging the limits of support from others who could not see the patient. Access to trauma and medical telephone advice/support varied between ambulance services, with the trauma desk often regarded as helpful with decisions about which hospital to attend (TU or MTU). ED staff noted that crews sometimes called them for advice, but considered that this support should be provided by the ambulance service. A lack of a clinical support/trauma desk in one ambulance service was regarded as a factor in pre-alert variation and ED calls for advice.

*‘Can request EOC consultants to call if they’re on shift. ‘They’re absolutely amazing…we’ve got something called the Clinical Assessment Team. Now, they are paramedics. So, for me, are of no clinical benefit because it’s somebody of the same grade*.*’ [AS9_Paramedic]*

*‘If we’re somewhere else on the patch and we’re not sure who can deal with what, we’ll ring up the trauma desk*.*’ [AS20_Paramedic]*

*‘It’s time critical I need this advice now…they [clinical support] just don’t answer the phone’ [AS27_Paramedic]*

*‘It does seem that [Ambulance service] has a lower level of qualification, generally, there are a lot more techs than paramedics, and they don’t have so much central support. They don’t have a trauma desk…they seem overly reliant on ED staff*.*’ [ED46_Consultant]*

Colleagues were also mentioned as a potentially valuable source of support, depending on their level of experience and qualification. More experienced/specialist staff could be called to attend where necessary. The hospital-based HALO also sometimes acted as a helpful intermediary, passing on information and advocating for crew decisions.

*‘I’ll always talk it [decision] through…any of my colleagues input is just as important, doesn’t mean I’m going to do what they think but just getting their input can help my decision-making*.*’ [AS43_ Emergency Medical Technician]*

### Consideration of differences between receiving Emergency Departments

Differences in pre-alert criteria between ambulance services and EDs created difficulties in deciding whether to pre-alert and was particularly challenging for less experienced clinicians and when pre-alerting an out-of-area ED.

*‘Each hospital seems to have a different level of what they would like to be pre-alerted… that makes the decision making difficult*.*’ [AS4_Paramedic]*

*‘When you work at [Ambulance Service], then you’ve got 6/7 different hospitals that all work differently…phone numbers change, policies change and its generally word of mouth with each other that we get it passed round*.*’ [AS19_Paramedic]*

*‘If we go 6 miles further south from our location, we’ve got a choice of two hospitals that we go to… both of them are completely out of area and have local protocols where they’ll change things and people in the area will know but people from out the area don’t necessarily know*.*’ [AS20_Paramedic]*

Ambulance Service protocols were generally perceived to have lower thresholds for triggering pre-alerts, especially for children and suspected stroke or sepsis, which was attributed to organisational risk aversion.

*‘What makes the pre-alert decision hard as a clinician is if our policy and protocols are saying it’s for pre-alert, but they don’t marry up to the hospitals expectations of what they’d accept in resus*.*’ [AS39_Clinical Lead]*

*‘All strokes have to be alerted whether they are in time or not. That is definitely over-alerting it. Children and children that are alerted I think because again, their [ambulance service] markers are different to ours… when they’ve inputted their obs. it can automatically trigger that they need to alert the patient*.*’ [ED15_Senior Nurse]*

Frustration was expressed by both services regarding the sepsis protocols as crews feel obliged to alert due to the low threshold, and whilst EDs recognised this to be the case, ‘protocol-driven’ pre-alerts could be a source of tension, particularly when very busy.

*‘Sepsis pre-alerts are always ones that you might be umm-ing and ahh-ing because with sepsis, the sepsis inclusion criteria are quite low for the threshold*.*’ [AS37_Paramedic]*

*‘Our [ambulance service] interpretation of sepsis is far different than an ED*.*’ [AS20_Paramedic]*

When ambulance and ED pre-alert criteria differed, ambulance clinicians experienced anxiety about the reaction they would receive. Similarly, a degree of hesitancy was experienced when deciding to pre-alert hospitals perceived as generally less receptive to pre-alerts. Decisions about which hospital to attend were sometimes more difficult than identifying that a pre-alert was needed due to variations in criteria, especially whether the patient needs to go to a specialist centre (e.g. MTC).

*‘Some of my hospital colleagues, I also don’t think they really understand the context in which we work, and the fact that we don’t always get it right. But actually the best hospitals recognise that, accept what we’re saying*.*’ [AS29_ Specialist Paramedic]*

Delays at receiving hospitals were a consideration where there was a choice of hospitals to attend and concerns over potential deterioration if they had to wait. ED staff recognised that a reduced threshold for pre-alerting was inevitable due to delays.

*‘Would I want this patient sat in a corridor for a little while or not? And if the answer’s no then I pre-alert it’ [AS31_Paramedic]*

*‘We can be waiting one, two hours potentially. So if you know it’s a busy one at the hospital then I might be more inclined to pre-alert just so then the hospital staff have got the opportunity, if they want to, to potentially jiggle things around in the HDU and make some room… Obviously once we get there it can be downgraded*.*’ [AS31_Paramedic]*

*‘[If] we’re equidistant between the 2 hospitals, we’ll always pick the one with the lower queue’ [AS19_Paramedic]*

Clinicians reported not making pre-alert calls for time-critical patients when close to the hospital and when alternative mechanisms in place enabled them to ask resus for an assessment on arrival. Some concern was expressed about pre-alerts being used to circumvent queues, but many commented that this view was unwarranted, with ambulance clinicians emphasising that doing so would undermine trust in their pre-alerts.

*‘A lot of the time it’s to do with distance, so they’ll say well we were only five minutes around the corner…but it still would have given me four or five minutes to set things up for this patient, and that is part of their protocol so it’s a bit unacceptable really, but again I always speak to the crews*.*’ [ED33_ Senior Nurse]*

*‘pre-alerting doesn’t necessarily improve your chances of not waiting in overflow, so I wouldn’t say it gets used that way*.*’ [AS1_Emergency Medical Technician]*

*‘I’ve not met a crew that have alerted a patient to finish on time’ [ED44_Nurse]*

## DISCUSSION

### Summary of findings

The findings highlight a range of individual and organisational level factors potentially contributing to variation in pre-alert decision-making. Different levels of experience and confidence affected ambulance clinician judgments about which patients required immediate ED attention or were at greatest risk of deterioration. Understanding how to pre-alert involved learning ‘on the job’ and informal feedback, including assumptions based on the ED response to previous pre-alerts. Clinical decision support varied between services, including access to clinical advice, and the pre-alert criteria specified in guidance and protocols. Differences in pre-alert criteria between ambulance services and EDs created difficulties in deciding whether to pre-alert, particularly for less experienced clinicians. More experienced clinicians tended to balance the use of decision support with autonomous clinical judgement.

### Relevant research

Previous research has identified the benefits of pre-alerts for patient care and outcomes^3 4^ but variation in practice has also been identified, even for clinical conditions acknowledged as clearly necessitating a pre-alert.^7 16^ The most recent evidence in relation to pre-alert practice across a range of clinical conditions is from studies designed to complement the research in this paper.^6 17 18^ The findings presented here provide insights into potential sources of variation in pre-alert practice that may explain variation in pre-alert rates (8%-15%) between the same three services.^17^

At an individual level, newly qualified clinicians were identified as being more reliant on decision support and generally more risk averse in pre-alert decisions, which is consistent with evidence indicating that newly qualified paramedics pre-alert more than experienced clinicians across the three services.^17^ A survey of ambulance clinicians across all 10 services in England^18^ found differences in pre-alert assessment practices, sources of decision support and understandings of conditions that need pre-alerting, even for conditions where pre-alerts would be expected for all patients. Accounts by both ambulance and ED clinicians of variation in pre-alert criteria is consistent with findings from research by Boyd et al. ^6^ that identified differences between national and local pre-alert guidance, as well as inconsistencies in pre-alert thresholds for a range of conditions. Paramedic discretion in pre-alert decisions has also been shown to play a significant role in identifying patients at high risk who may not be captured by pre-alert guidance and decision tools.^19^

A review of ambulance clinician decision-making explains experience-related variation as reflecting the ability of expert clinicians to draw on more extensive experiences to inform conscious and subconscious thought processes involved in forming rapid clinical impressions of critically ill patients.^20^ The authors highlighted the importance of routine reflection and feedback for continued improvement. Wilson et al.^21^ highlight the potential benefits of feedback for clinical practice, patient outcomes and staff mental health. In common with the findings from this study, they identify lack of feedback as an issue for ambulance clinicians in the UK and internationally, despite an expressed desire for such information.

Research exploring ambulance clinician non-conveyance decisions, similarly identified that risk averseness was influenced by levels of experience, confidence, and perceptions of vulnerability if something goes wrong.^22^ Ambulance clinician accounts that the perceived risk of deterioration was amplified by anticipated ED delays was consistent with findings from quantitative analysis of pre-alert practice in the three participating services, which showed pre-alerts increased when there were delays at EDs.^17^ Sujan et al.^23^ identified this lowering of the threshold for pre-alert decisions due to delays as potentially diverting resources away from where they are most needed if the pre-alerts are not necessary. The potential burden on ED resources and staff of decisions to pre-alert when unnecessary has been identified in other studies.^5 7 16^

### Limitations

Observation at the point of pre-alert decision-making could have enhanced the study findings but was not feasible due to the relatively small number of pre-alerts occurring in each shift. We originally aimed to compare clinicians with high and low rates of pre-alerts but were unable to obtain the necessary routine data in time. Although the number of interviewees reflected a pragmatic decision, analysis indicated thematic saturation. Triangulation involving multiple researchers and data sources, was designed to enhance the rigour and trustworthiness of the research.^24^ This research involved only three ambulance services but a linked survey of all ambulance services in England provides complementary evidence from a larger sample of services and clinicians.^18^

Further research could examine the impact of changes to guidance, processes, training and feedback, including what type of feedback is most effective. Research looking at pre-alert practice could explore differences between clinicians with different pre-alert rates and possibly levels of experience.

### Implications

We identified potentially avoidable variation in pre-alert decision-making, which has implications for patient care and emergency care resources, and can contribute to tension between the services. By providing insight on individual and organisational level variation in pre-alert decision-making, the study highlights areas for attention to develop more consistent pre-alert practice and patient care. Consistency in practice may be improved by greater standardisation of guidance and protocols, training and access to performance feedback, and cross-service collaboration to minimise potential sources of tension. The balance between adherence to standardised pre-alert criteria and accommodating autonomous clinical judgement also merits consideration.

## Data Availability

The data generated for this study is in the form of confidential transcripts of interviews that are not available for sharing. Participants consented for anonymised quotations to be shared but did not consent to share the full transcripts.

## Funding statement

This research was funded by the National Institute for Health and Care Research (NIHR HS&DR 131293). The views expressed in this publication are those of the author(s) and not necessarily those of the NIHR or the UK Department of Health and Social Care.

The views expressed are those of the author(s) and not necessarily those of the NIHR or the Department of Health and Social Care.

## Acknowledgements

The authors would like to thank all research participants and ED and ambulance service staff who helped to recruit participants. We are also grateful for the input of other members of the study team, our advisory group and our patient and public involvement representatives/group. Thanks to Marc Chattle for clerical support.

## Notes

### Competing Interest Statement

The authors have declared no competing interest.

### Author Declarations

NHS ethics North East - Newcastle & North Tyneside 2 Research Ethics Committee (Ref: 21/NE/0132) gave ethical approval for this work

